# Wastewater based epidemiology beyond SARS-CoV-2: Spanish wastewater reveals the current spread of Monkeypox virus

**DOI:** 10.1101/2022.09.19.22280084

**Authors:** Inés Girón-Guzmán, Azahara Díaz-Reolid, Pilar Truchado, Albert Carcereny, David Garcia-Pedemonte, Bruno Hernaez, Albert Bosch, Rosa María Pintó, Susana Guix, Ana Allende, Antonio Alcamí, Alba Pérez-Cataluña, Gloria Sánchez

**Affiliations:** VISAFELab, Department of Preservation and Food Safety Technologies, Institute of Agrochemistry and Food Technology, IATA-CSIC, Av. Agustín Escardino 7, Paterna, Valencia 46980, Spain; Department of Food Science and Technology, CEBAS-CSIC, Research Group on Quality and Safety of Fruits and Vegetables, Campus Universitario de Espinardo, 25, Murcia 30100, Spain; Enteric Virus Laboratory, Department of Genetics, Microbiology, and Statistics, Section of Microbiology, Virology, and Biotechnology, School of Biology, University of Barcelona, Barcelona, Spain; Molecular Biology Center Severo Ochoa, CSIC-UAM, Campus de Cantoblanco, Nicolás Cabrera 1, 28049 Madrid, Spain

**Keywords:** monkeypox, wastewater, epidemiology, WBE

## Abstract

Besides nasopharyngeal swabs, monkeypox virus (MPXV) DNA has been detected in a variety of samples such as saliva, semen, urine and fecal samples. Using the environmental surveillance network previously developed in Spain for the routine wastewater surveillance of SARS-CoV-2 (VATar COVID-19), we have analyzed the presence of MPXV DNA in wastewater from different areas of Spain. Samples (n=312) from 24 different wastewater treatment plants were obtained between May 9 (week 22_19) and August 4 (week 22_31), 2022. Following concentration of viral particles by flocculation, a qPCR procedure allowed us to detect MPXV DNA in 63 wastewater samples collected from May 16 to August 4, 2022, with values ranging between 2.2 × 10^3^ to 8.7 × 10^4^ genome copies (gc)/L. This study shows that MPXV DNA can be reproducibly detected by qPCR in longitudinal samples collected from different Spanish wastewater treatment plants. According to data from the National Epidemiological Surveillance Network (RENAVE) in Spain a total of 6,119 cases have been confirmed as of August 19, 2022. However, and based on the wastewater data, the reported clinical cases seem to be underestimated and asymptomatic infections may be more frequent than expected.

## 1. Introduction

In early May 2022, a multi-country outbreak of Monkeypox virus (MPXV) started in non-endemic regions, and on 23 July WHO declared a Public Health emergency of international concern (WHO, 2022). In Europe, a total of 13,911 cases of MPX have been reported up to 19 August 2022, with Spain accounting for 6,119 cases, the second highest number of monkeypox (MPX) cases worldwide, being present in most regions of the country (Spanish Ministry of Health, 2022).

Symptoms developed include the appearance of rash, fever, fatigue, muscle pain, vomiting, diarrhea, chills, sore throat or headache, and the hospitalization rate is around 8-13% (European Centre for Disease Prevention and Control (ECDC), 2022.; Thornhill et al., 2022). Sadly, two deaths linked to this outbreak have occurred in Spain due to complications associated with encephalitis (Aguilera-Alonso et al., 2022). It is assumed that transmission occurs after close contact with skin lesions of an infected person, as well as through contact with respiratory droplets and fomites, and that infection is symptomatic in all patients (McCollum and Damon, 2014). However, antibodies have been found in exposed asymptomatic individuals, which can be linked to subclinical infections (Wilson et al., 2014), and positive MPXV PCR results from anal samples in asymptomatic men who have sex with men (MSM) have also been documented (Baetselier et al., 2022; Ferré et al., 2022). The virus is also excreted in fluids, and its detection in saliva, semen, urine and feces has been reported (Peiró-Mestres et al., 2022). This implies that routine wastewater surveillance can be applied as a tool for early detection of the disease expansion as very recently reported according a model-based theoretical evaluation (Chen and Bibby, 2022). According with this model, wastewater-based epidemiology (WBE) can detect on average 7 MPX cases out of 100,000 people. Currently, various studies detected MPXV DNA in wastewater worldwide (de Jonge et al., 2022; la Rosa et al., 2022; Wolfe et al., 2022), highlighting again wastewater analysis as a non-invasive tool for monitoring the status and trend an emerging infection. The aim of the present study was to trace the community circulation of the MPXV from potentially symptomatic, asymptomatic, or presymptomatic individuals using the previous established Spanish National SARS-CoV-2 Wastewater Surveillance Network (VATar COVID-19).

## 2. Material and methods

### 2.1. Sample concentration and DNA extraction

Grab sewage samples were collected from 24 Spanish wastewater treatment plants (WWTPs) (Fig. 1C) between May 9 (week 22_19) and August 4 (week 22_31) and kept at 4°C until analysis. Concentration of viral fraction was performed with a previously validated method for SARS-CoV-2 using an aluminum-based adsorption precipitation procedure (Pérez-Cataluña et al., 2021). Nucleic acids extraction of the concentrated samples was performed with the Maxwell® RSC Instrument (Promega) using the Maxwell RSC Pure Food GMO and authentication kit (Promega) and the “Maxwell RSC Viral total Nucleic Acid” program.

**Figure 1.**
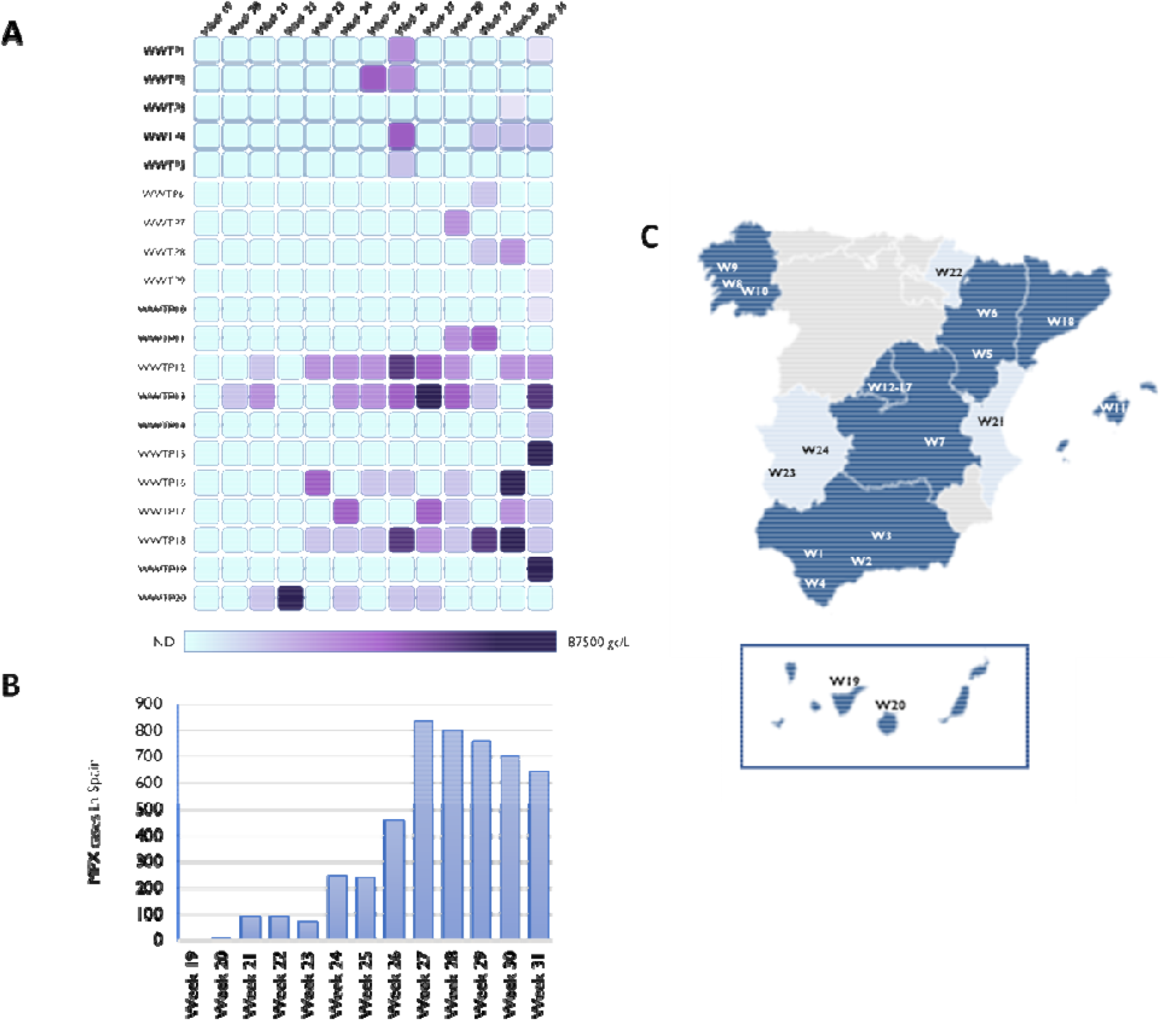
**A)** Evolution of MPXV DNA prevalence over time, as measured by qPCR in wastewater samples from 20 wastewater treatment plants with positive detection **B)** Number of cases of monkeypox per week (Spanish Ministry of Health) **C)** Geographical localization of wastewater treatment plants, dark blue (Autonomous Community with positive detection in the analyzed wastewater samples) and light blue (no detection in wastewater samples).

### 2.2. MPXV real -time PCR assays

The MPXV West Africa (G2R_WA) assay (Li et al., 2010) was used to quantify MPXV DNA using the qPCR Premix Ex Taq™ kit (Takara Bio Inc). Additionally, a subset of samples (Table S1) was tested for MPXV DNA using the MPXV generic (G2R_G) assay (Li et al., 2010). Undiluted and ten-fold diluted DNA was tested in duplicated. Positive control consisted in the nucleic acid material extracted from a cell culture infected with a clinical MPXV specimen obtained from a patient pustule. For each qPCR, serial dilutions of standard curves were run in quintuplicates and the numbers of estimated genome copies were calculated (Table S2). Each run included negative controls (nuclease-free water and negative extraction controls). Depending on the laboratory, reactions were carried out in the QuantStudio™ 3 and QuantStudio™ 5 Real-Time PCR Systems (ThermoFisher Sci.)

## 3. Results

### 3.1 Estimated levels of monkeypox virus DNA in wastewater samples

Here, we report the first detection of MPXV DNA in wastewater samples from different regions of Spain. 63 out of 312 samples showed positive results for MPXV DNA, corresponding to samples collected from week 22_20 to week 22_31. Cycle threshold values ranged between 44.3 and 34.5, corresponding to values from 2.2 × 10^3^ to 8.7 × 10^4^ estimated genome copies (gc) per liter (Fig. 1A). First detection of MPXV DNA in wastewater samples occurred in WWTP13 from the city of Madrid in week 22_20 (Fig. 1A and Fig. 2), with positive detection using two different assays (Table S1). On that week, Madrid reported the first suspected cases of MPX which represented the first cases of MPX in Spain accounting for one of the largest outbreaks reported outside Africa (Iñigo Martínez et al., 2022). Later on, several cases were reported in Madrid before the outbreak declaration on 17 May, most of them attending the same sauna in the city of Madrid or with travel history to Maspalomas Gay Pride festival that took place on 5-15 May in Gran Canaria.

**Figure 2.**
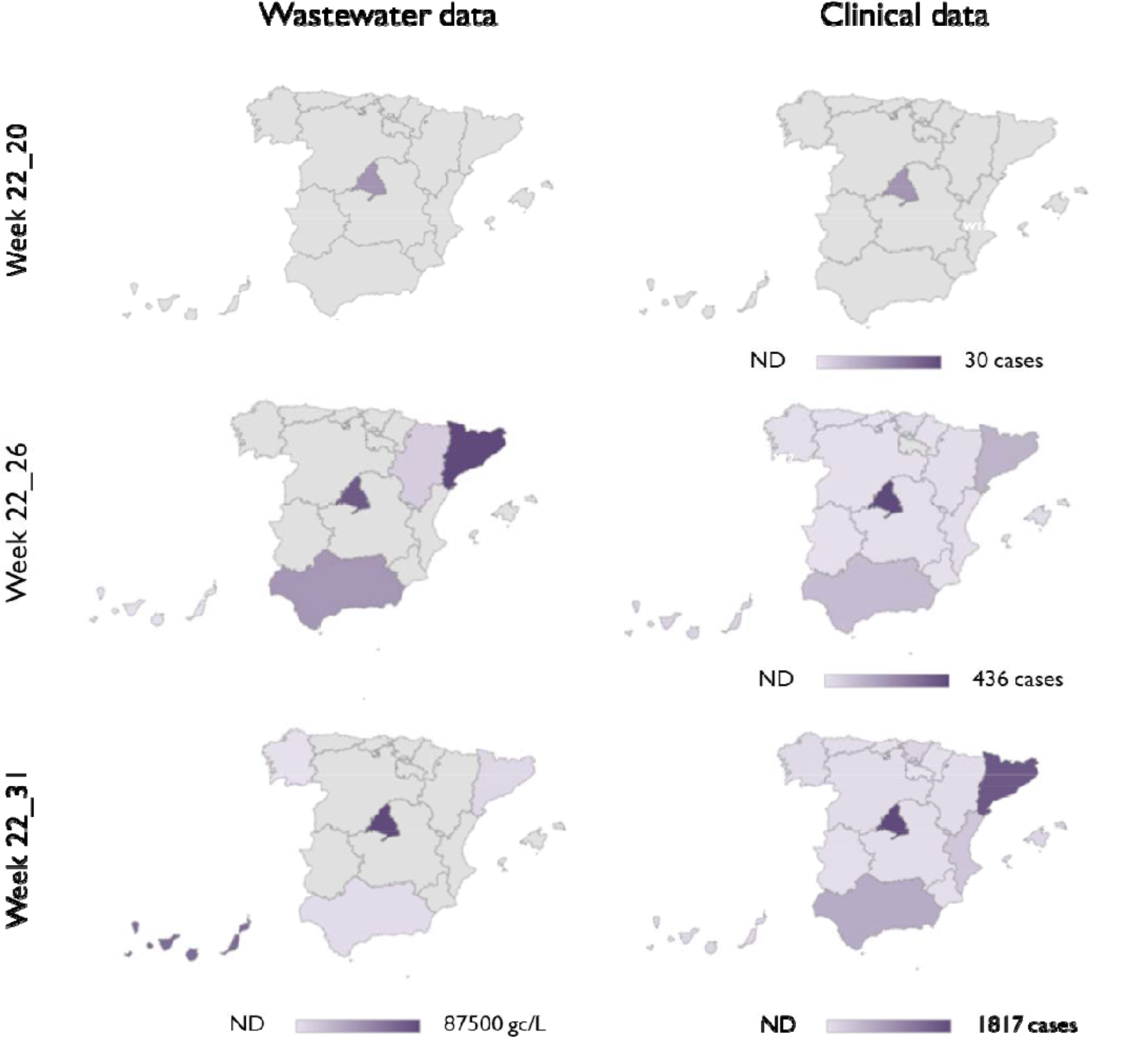
Comparison of MPXV DNA estimates from wastewater testing (left panels) and confirmed cases of monkeypox by Autonomous community reported by the Health Ministry authorities (right panels). For wastewater samples, highest level within the same Autonomous Community are depicted. ND: No detection

In week 22_21, MPXV DNA was detected in the closest WWTPs of Madrid city (WWTP12 and WWTP13) and in WWTP20 of Gran Canaria (Canary Islands) (Fig. 1, Table S1). Interestingly, we consistently detected MPXV DNA in samples collected from week 22_23 in WWTP12 and WWTP13 from Madrid, when only 275 cumulative cases were declared in the entire Region (Iñigo Martínez et al., 2022). Our data also showed percentages of WWTPs with MPXV DNA detection increased progressively, up to 15% by week 22_23 and 45% by week 22_26 (Fig. 1A and Fig. 2). In Barcelona, the second largest Spanish city, first detection occurred in week 22_23 in WWTP18 with the first peak observed in week 22_26 when 130 cumulative cases were detected in Catalonia (Fig. 2). Intermittent detection (negative results after previous qPCR detection) was reported from some WWTPs where the number of confirmed clinical cases was low (Fig. 1 and Fig. 2). Furthermore, all weekly samples collected from Valencia (WWTP21), Extremadura (WWTP23 and WWTP24), and Navarra (WWTP22) tested negative for the presence of MPXV DNA (Fig. 2), with a total number of clinical cases of 331, 21 and 13 as August 9, respectively (Spanish Ministry of Health, 2022).

## 4. Discussion

The COVID-19 pandemic has demonstrated that WBE is a cost-effective tool to anticipate the circulation of SARS-CoV-2 in a community and to closely track its incidence, evolution and geographic spread (Bivins et al., 2020). WBE has been implemented worldwide and most of the countries are ready to perform this monitoring as a routing basis for other emerging pathogens likely to be found in wastewater, due to their presence in feces and/or urine. In Spain, the National WBE Network, VATar COVID-19, has been successfully used to determine the extent of the COVID-19 disease along the country (Carcereny et al., 2021).

The increasing number of MPXV cases around the world continue to pose challenges to control its transmission with a total number of 41,358 cases as of 19 Aug 2022 (CDC, 2022). This underscores the urgent needs for simple and cost-effective tools to facilitate early detection, evolution and spatial distribution of cases. DNA of MPXV has been detected in urine and feces from symptomatic individuals (Antinori et al., 2022; Peiró-Mestres et al., 2022), and although limited data are available, viral shedding has been observed in stool in 63 % of patients (Cts values from 17.8 to 31.4) and in urine in 56% (Cts values from 19.1 to 40.0) (Peiró-Mestres et al., 2022). It is not known whether MPXV present in stool and urine is infectious. Altogether, these findings warned the interest of assessing the presence of MPXV DNA in sewage samples (Chen and Bibby, 2022). In the current study, a qPCR assay designed for the West African clade (Li et al., 2010) was applied on wastewater samples collected from week 22_19 to week 22_31, showing that MPXV DNA can be reproducibly detected by qPCR in longitudinal samples collected from several Spanish WWTPs. First detection of MPXV DNA was retrieved in a single sample from WWTP13 collected on May 17 (Week 22_20) using the specific qPCR assay and confirmed by the MPXV generic assay, providing the earliest piece of evidence that the virus was circulating in the community of Madrid. Interestingly, we consistently detected MPXV DNA in samples collected in WWTP18 (Barcelona) since week 22_23, when only 39 cumulative cases were declared in the entire Autonomous Community of Catalonia. In line, MPXV DNA was also detected in week 22_21 on wastewater samples collected from Schiphol Airport and in different Dutch WWTPs from week 22_22 onwards (de Jonge et al., 2022).

The viral concentration method used in this study has been validated for SARS-CoV-2 detection and quantification (Pérez-Cataluña et al., 2021) and it seems promising for MPXV monitoring in wastewater, too. However, in contrast to what has been reported for SARS-CoV-2 (Bivins et al., 2020; Medema et al., 2020; Randazzo et al., 2020), anticipation has not been observed for MPXV, for which the first wastewater detection occurred at the same time that MPXV cases were declared (Fig. 2). This could be due to several factors, including differences in shedding levels and kinetics, proportion of asymptomatic cases, diagnosis of the disease and fast identification of cases, environmental factors affecting virus stability, low performance of the method to concentrate MPXV, and also a much larger scale of transmission of SARS-CoV-2 in the community. Therefore, further assessment of the performance characteristics of the methodology needs to be carried out. However, it is important to highlight that wastewater positive samples have been found in areas with very low reported disease prevalence. For instance, in Castilla la Mancha, a region located at the middle-south of Spain, MPXV was detected in sewage with only 42 clinical cases being reported, indicating that probably, a higher number of people may be affected. As previously discussed by other authors, stigma and discrimination may be limiting the awareness or willingness of at-risk people to have their symptoms evaluated. In these situations, WBE may be even more useful, because the anonymous pooled samples can evidence the contributions of a community without divulging individual identities (Nelson, 2022).

## 5. Conclusions

Using an environmental surveillance tool previously developed for SARS-CoV-2, we have been able to consistently detect MPXV DNA in wastewater samples from different regions of Spain when communicated clinical cases in that region were only incipient. We also found that the wastewater viral DNA detection increased rapidly and anticipated the subsequent ascent in the number of declared cases showing, once again, that WBE is a sensitive and cost-effective strategy for the surveillance emerging viral threats. In those cases where stigma and blame might undermine the capacity to effectively respond during outbreaks, i.e., driving people away from health services, the implementation of WBE may represent a most valuable tool.

## Data Availability

All data produced in the present work are contained in the manuscript

## Acknowledgements

This research was supported by the European Commission NextGenerationEU fund, through CSIC’s Global Health Platform (PTI Salud Global) and samples were obtained from the COVID-19 wastewater surveillance project (VATar COVID-19) funded by the Spanish Ministry for the Ecological Transition and the Demographic Challenge and the Spanish Ministry of Health. IGG is recipient of a predoctoral contract from the Generalitat Valenciana (ACIF/2021/181) and AP-C was supported by a postdoctoral fellowship (APOSTD/2021/292). PT is holding a Ramón y Cajal contract from the Ministerio de Ciencia e Innovación and AC is recipient of a predoctoral contract FI-SDUR from the Generalitat de Catalunya.

