## Supplementary for "Wastewater based epidemiology beyond SARS-CoV-2: Spanish wastewater reveals the current spread of Monkeypox virus"

**Table S1.** Monkeypox virus DNA detection in Spanish wastewater samples.

|  |  | **G2R_WA** | | | | **G2R_G** | |
| --- | --- | --- | --- | --- | --- | --- | --- |
|  |  | **Undiluted DNA** | | **Ten-fold diluted DNA** | | **Undiluted DNA** | |
| **WWTP** | **Week** | **Ct value 1** | **Ct value 2** | **Ct value 1** | **Ct value 2** | **Ct value 1** | **Ct value 2** |
| WWTP1 | 26 | 36.71 | nd | nd | nd | 36.89 | nd |
| WWTP1 | 31 | 39.63 | nd | nd | nd | np | np |
| WWTP2 | 25 | 36.32 | nd | nd | nd | 37.21 | nd |
| WWTP2 | 26 | 36.59 | nd | nd | nd | 37.72 | nd |
| WWTP3 | 30 | 39.98 | nd | nd | nd | nd | nd |
| WWTP4 | 26 | nd | nd | nd | 39.77 | np | np |
| WWTP4 | 29 | 37.60 | nd | nd | nd | np | np |
| WWTP4 | 30 | nd | 38.1 | nd | nd | np | np |
| WWTP4 | 31 | 37.91 | nd | nd | nd | np | np |
| WWTP5 | 26 | 37.82 | nd | nd | nd | nd | nd |
| WWTP6 | 29 | 38.76 | nd | nd | nd | np | np |
| WWTP7 | 28 | 37.09 | nd | nd | nd | nd | nd |
| WWTP8 | 29 | 37.86 | nd | nd | nd | nd | nd |
| WWTP8 | 30 | 36.86 | nd | nd | nd | nd | nd |
| WWTP9 | 31 | 39.56 | nd | nd | nd | np | np |
| WWTP10 | 31 | 39.65 | nd | nd | nd | np | np |
| WWTP11 | 28 | 39.98 | nd | nd | nd | nd | nd |
| WWTP11 | 29 | 36.76 | nd | nd | nd | 38.61 | nd |
| WWTP12 | 21 | 37.74 | nd | nd | nd | nd | nd |
| WWTP12 | 23 | 36.91 | nd | nd | nd | nd | nd |
| WWTP12 | 24 | 36.90 | nd | nd | nd | 37.86 | nd |
| WWTP12 | 25 | 36.89 | nd | nd | nd | 36.73 | nd |
| WWTP12 | 26 | 35.47 | nd | nd | nd | 37.24 | nd |
| WWTP12 | 27 | 36.52 | nd | nd | nd | 39.81 | nd |
| WWTP12 | 28 | 36.84 | 43.69* | nd | nd | 37.18 | 36.42 |
| WWTP12 | 30 | 39.90 | 37.83 | nd | nd | np | np |
| WWTP12 | 31 | 36.99 | nd | nd | nd | np | np |
| WWTP13 | 20 | 37.61 | 42.75* | nd | nd | 38.26 | nd |
| WWTP13 | 21 | 36.76 | nd | nd | nd | 38.90 | nd |
| WWTP13 | 22 | nd | nd | nd | nd | 38.18 | nd |
| WWTP13 | 23 | nd | nd | nd | nd | 38.56 | nd |
| WWTP13 | 24 | 40.94* | 36.03 | nd | nd | nd | nd |
| WWTP13 | 25 | 36.82 | nd | nd | nd | 38.37 | 36.84 |
| WWTP13 | 26 | 36.17 | 36.24 | nd | nd | 37.22 | 37.55 |
| WWTP13 | 27 | 34.50 | 35.02 | nd | nd | 36.03 | 36.90 |
| WWTP13 | 28 | 35.93 | 36.76 | nd | nd | 36.09 | 36.87 |
| WWTP13 | 29 | 37.78 | nd | nd | nd | 38.49 | nd |
| WWTP13 | 31 | 36.94 | 34.94 | nd | nd | np | np |
| WWTP14 | 31 | 37.44 | nd | nd | nd | np | np |
| WWTP15 | 31 | 34.71 | 34.33 | nd | nd | np | np |
| WWTP16 | 23 | nd | nd | 39.16 | nd | np | np |
| WWTP16 | 25 | 40.62* | nd | nd | nd | np | np |
| WWTP16 | 26 | 40.57* | nd | nd | nd | np | np |
| WWTP16 | 28 | 39.89 | 40.65* | nd | nd | np | np |
| WWTP16 | 30 | 38.32 | 37.43 | nd | 37.23 | np | np |
| WWTP17 | 24 | 40.53* | nd | 40.06* | nd | np | np |
| WWTP17 | 27 | 40.77* | 39.08 | nd | 39.97 | np | np |
| WWTP17 | 28 | 42.28* | nd | nd | nd | np | np |
| WWTP17 | 30 | 36.91 | 37.22 | nd | nd | np | np |
| WWTP17 | 31 | 37.78 | nd | nd | nd | np | np |
| WWTP18 | 23 | 41.16* | nd | nd | nd | np | np |
| WWTP18 | 24 | 37.85 | nd | nd | nd | np | np |
| WWTP18 | 25 | 37.83 | 38.3 | nd | nd | np | np |
| WWTP18 | 26 | 39.49 | 37.85 | 38.37 | 44.3* | np | np |
| WWTP18 | 27 | 37.52 | 37.99 | nd | nd | np | np |
| WWTP18 | 28 | nd | 39.04 | nd | nd | np | np |
| WWTP18 | 29 | 36.74 | 38.57 | 37.82 | nd | np | np |
| WWTP18 | 30 | nd | 38.18 | 37.77 | nd | np | np |
| WWTP18 | 31 | 37.77 | 38.95 | nd | nd | np | np |
| WWTP19 | 31 | nd | nd | 37.93 | nd | np | np |
| WWTP20 | 21 | 40.15* | nd | nd | nd | np | np |
| WWTP20 | 22 | nd | nd | nd | 37.6 | np | np |
| WWTP20 | 24 | 40.53* | nd | nd | nd | np | np |
| WWTP20 | 26 | 39.39 | nd | nd | nd | np | np |
| WWTP20 | 27 | nd | 39.05 | nd | nd | np | np |

Ct: cycle threshold; nd: non detected; np: non performed; *: Ct values above 40

**Table S2.** Parameters defining standard curves for of all 3 participating laboratories for the MPXV West Africa (G2R_WA) and generic (G2R_G) assays [1].

| **Laboratory** | **Target** | **Slope** | **Intercept** | **R2** |
| --- | --- | --- | --- | --- |
| **A** | G2R_WA | **-3.43** | **39.38** | **0.99** |
|  | G2R_G | **-3.31** | **40.16** | **0.99** |
| **B** | G2R_WA | **-3.29** | **39.50** | **0.98** |
| **C** | G2R_WA | **-3.46** | **39.37** | **0.99** |
